# Anti-retroviral therapy and serum protein levels in HIV-1 seropositive patients: a five-year retrospective study

**DOI:** 10.1101/2020.06.23.20129338

**Authors:** Akor Egbunu Shedrac, Musa Haruna, Eneojo-Abah Eleojo Gloria, Yisa Benjamin Nma, Emmanuel Friday Titus, Dickson Achimugu Musa, Joel Ikojo Oguche, Serah Shaibu, Salami Tijani, David Bukbuk, Samuel Eneọjọ Abah

## Abstract

**Background:** Serum proteins designated as liver function biomarkers are used to evaluate patients for hepatic dysfunction. Hepatic effect of Anti-Retroviral Therapy (ART) needs further studies in HIV mono-infected patients. In this study, clinically defined patient datasets were analysed for protein levels in HIV-1 mono-infected seropositive patients with and without ART.

**Materials and Methods:** Data were collected for the study groups, consisting of the control group and HIV-1 mono-infected seropositive patients with and without ART and were analysed statistically for differences among the groups. All subjects in the patient groups attended University of Maiduguri Teaching Hospital, Nigeria for a period of 5 years.

**Result:** The protein levels on initiation of ART were significantly higher than baseline levels (prior to ART). However, continuous use of ART for 5-year period did not induce any further significant change in protein levels. Receiver Operating Characteristic (ROC) curves shows that both Albumin (ALB) and Total protein (TP) levels discriminated among the study groups. The baseline levels of ALB in seropositive patients are significantly lower to levels on initiation of ART.

**Conclusion:** Continuous ART did not cause any further significant change in levels of liver function proteins than was observed on ART initiation. Hence, liver damage on continuous ART is not implied. Both ALB and TP levels could be important in HIV management of patients. Initiation of ART appears to elevate the low ALB level via a yet unknown mechanism and indicates possible role of ALB in ART mechanism of action.

## 1.0 Introduction

Human Immune deficiency Virus (HIV) continues to be a major global health issue and have claimed more than 32 million lives (WHO, 2019a) and approximately 37.9 million people are living with HIV as at the end of 2018. The prevalence of HIV among African adults (15-49 years) is 3-4 times higher in 2018 (WHO, 2019b). Increased HIV in Africa is associated with poor policies and social factors and includes exposure to risk situations and lack of effective prevention, testing and treatment services. About 1.7 Million new cases and 770, 000 deaths were reported in Africa in 2018(WHO, 2019a).

Human Immunodeficiency Virus (HIV) is a retroviridae with a unique enzyme, the reverse transcriptase which is RNA directed DNA polymerase. On entry into a target cell the viral RNA genome is reverse transcribed into double stranded DNA and incorporated into cellular DNA with the aid of host co-factors and viral integrase and then either become latent for a while and/or transcribed, producing new RNA genome and viral proteins to begin new replication cycle. The virus destroys CD4 helper T cell, entry into CD4+ T cell is with the help of viral envelop glycoprotein (gp) gp120 and gp41 and CD4+ cells receptors present on macrophages and lymphocytes, the CCR5 or CXCR4 chemokine receptors. It attacks CD4 cells and spread via cell to cell or via blood to infect new T-cells(Smith & Daniel, 2006; Zhang et al., 2015). Its continuous destruction of the host cells compromises the immune system (Bobat & Archary, 2017; Coffin & Swanstrom, 2013; Greene & Peterlin, 2002; Grossman, Meier-Schellersheim, Paul, & Picker, 2006; Lackner, Lederman, & Rodriguez, 2012; Maartens, Celum, & Lewin, 2014; Moir, Chun, & Fauci, 2011; Naif, 2013; Swanstrom & Coffin, 2012; Wilen, Tilton, & Doms, 2012). Acquired Immune-Deficiency Syndrome (AIDS) is typical when the CD4+ T cell count is below 200 cells/µL with the occurrence of opportunistic infections, for example respiratory tract infection or pneumocystis pneumonia. Initial infection is not characterized with obvious symptoms or ill-feeling at which stage individual looks apparently healthy with slight underlying changes in the immune system during which seroconversion occurs, a period spanning up to 3 months after which detection of HIV specific antibodies becomes possible. The disease progression to severe HIV disease and immunosuppression varies with individual and often progress somehow slowly(Cunningham et al., 2000; Naif, 2013).

At present, there is no cure for HIV and Anti-Retroviral Therapy (ART) has helped to both control the virus and prevent onward transmission to other people. About 16.3 million (70.02%) HIV subjects were on ART in Africa(WHO, 2019a). Within the sub-Saharan Africa, 62% of adult, 82% of pregnant and breastfeeding women and 54% of children living with HIV are reported as receiving lifelong antiretroviral therapy, ART(WHO, 2019a).

ART induces varying degree of hepatic toxicity with conflicting reports (Baynes et al., 2017; Chu et al., 2010; Kalyesubula et al., 2011; Lucien, Clement, Fon, Weledji, & Ndikvu, 2010; Teklay, 2013) It is kind of complicated to accurately access the degree of hepatic damage associated with ART therapy in HIV subjects. This is partly because other than ART, hepatic damage in HIV subjects could be induced by several factors such as the pathophysiology of the infection itself; co-infection with hepatitis, majorly Hepatitis B-Virus (HBV) and Hepatitis C-Virus (HCV); patient lifestyle such as alcohol consumption and previous history of liver damage (Babu, Suwansrinon, Bren, Badley, & Rizza, 2009; Collins et al., 2006; Hoffmann et al., 2007; Maida et al., 2006; Neuman, Schneider, Nanau, & Parry, 2012; Puoti et al., 2009; Rhodes, 2005). Hence, a more clinically defined sample data set are required in order to ascertain the level of hepatic dysfunction in HIV subjects receiving ART. Most studies so far has been focused on HIV subjects co-infected with viral hepatitis (Coughlan, Valentine, Ruderman, & Saha, 2014; Lacombe & Rockstroh, 2012; Ocama et al., 2008; Osakunor, Obirikorang, Fianu, Asare, & Dakorah, 2015; Price & Thio, 2010; Thio & Locarnini, 2007; Trépo, Chan, & Lok, 2014). As a consequence, the relationship between HIV-1 mono-infected subjects on ART and associated hepatic damage remained largely unclear.

This study used a well clinically defined 5-year retrospective data set from HIV seropositive subjects with and without ART alongside with both their baseline and control data set, which are void of all possible cofounding factors. The data consist of serum protein levels, which are mostly markers of hepatic damage and measured in a cohort of well clinically defined HIV-1 mono-infected patients with and without ART and attending University of Maiduguri Teaching Hospital, Nigeria as well as the control group. The study accesses the impact of ART, the likely pathophysiology in subjects receiving ART and those not on ART as reflected on the serum protein level as well as the potential of these serum proteins in discriminating between the study groups.

## 2.0 Materials and Methods

### 2.1 Ethical Clearance and Informed Consent

The Research and Ethical Committee at the University of Maiduguri Teaching Hospital (UMTH), Nigeria, reviewed the study and gave a written ethical approval for the sample data collection at the Immunology department within the teaching hospital. Data were extracted from medical records in HIV clinics at the UMTH based on patient’s history. The study participants gave informed consent in accordance with the World Medical Association’s ethical principles for research involving human subjects.

### 2.2 Subject and Case Definition

A total of 150 subject data (Table 1) were studied and include 100 subjects with well clinically defined seropositive HIV status, out of which 50 were on ART for 5 years and are designated in this study as group A and 50 were yet to commence ART as at the time of diagnosis and sample collection and are designated in this study as group C. The control group consist of 50 healthy subjects without HIV infection and any other overt aetiology and are designated in this study as group B. All seropositive patients are diagnosed with HIV-1 infection.

**Table 1.** The cohort summary table.

| Variables | HIV +ve with ART<br>(Group A) | HIV -ve<br>Healthy group<br>(group B) | HIV +ve<br>without ART<br>(Group C) |
| --- | --- | --- | --- |
| No. | 50 | 50 | 50 |
| SEX (F/M) | 24/26 | 26/24 | 24/26 |
| AGE (Median, IQR) | 33.5(27-59) | *24(21.75-60) | 35.5(29.5-59) |
| DD (Median, IQR) | 3 (2-4.25) | N/A | N/A |
| Baseline data | Present | N/A | N/A |
| CD4 Counts (Range;<br>cells/μL) | 650-750 | 800-1200 | 320-450 |

**Notes:** \* statistically different ( $p = 0.001$ ).
**Abbreviations:** ART, anti-retroviral therapy; F/M, female/male; IQR, interquartile range. No, number; -ve, negative; +ve, positive; N/A, not applicable; HIV, human immunodeficiency virus

Subjects with diagnosed liver damage and co-infection with hepatitis (HBV and HCV) were excluded from the study. Alcoholic patients and those on drug with potential effect on the liver and those with abnormally high serum profile not attributable to the disease and indicative of pre-existing liver damage were removed from the study. All subjects yet to commence antiretroviral therapy were placed on the drug following diagnosis and sample collection.

The immune status based on CD4 counts (Table 1) are above the levels associated with AIDs and opportunistic infections. The study groups have no additional overt aetiology; however, clinical record shows they were still treated against opportunistic infections. CD4 counts in the disease groups (Group A and C) ranges from 320-750 cells/µL while the control group has CD4 counts in the range of 800-1200 cells/µL (Table 1).

Both males and females were equally represented in the study (Table 1). The age of the study participants is within the same range except that the HIV negative subjects have a slightly lower median that is significantly different (p = 0.001). This is unlikely to affect the result as the proteins level data that were analysed in the study are independent of age.

For this study, ART was defined as combined use of three or more antiretroviral drugs/regimen recommended for adults within the first line; TDF, tenofovir + 3TC, lamivudine (or FTC, emtricitabine) + EFV, efavirenz or AZT, zidovudine + 3TC + EFV (or NVP, nevirapine) or TDF + 3TC (or FTC) + DTG, dolutegravir or TDF + 3TC (or FTC) + EFV or TDF +3TC (or FTC) + NVP in accordance with the WHO guidelines on the use of antiretroviral drugs for treating and preventing HIV infection(WHO, 2016).

### 2.3 Assay procedure and data collection

The assay procedure leading to the collection of data analysed is here described. Venous blood (5ml) were collected from each subject, dispensed into plain sample container, allowed to clot and retract. The serum used for the analyses were separated from the clotted blood after centrifugation (Hettic centrifuge D-78532, Germany). Serum was screened for HIV-1/HIV-2 with OralQuick ADVANCE Rapid HIV-1/HIV-2. All positive subjects were diagnosed with HIV-1. A further HIV-1 Western Blot (Cambridge Biotech Corp) was performed to validate the assay. A western blot assay was considered negative if no band were present and positive if at least any two band from either the protein from the viral envelop (gp41, gp120/gp160); viral core (p17, p24, p55) and enzymes that are implicated in the pathophysiological process (p31, p51 and p66) were present. Subjects with inconclusive western blot were encouraged to repeat the test after a period of 3-4 weeks and were included in the study if assay show to be positive subsequently.

Serum were screened for HBV and HCV and positive sample data removed from the study. Briefly, HBV profile and anti-HCV antibody testing were done using immunochromatographic techniques to identify HBV surface antibody (HBsAb), HBV surface antigen (HBsAg), HBV e antigen (HBVeAg) and HBV e antibody (HBVeAb) and the core antibody (HBcAb) using a test kits and the manufacturer’s instructions (Guangzhou Wondfo Biotech Co. Ltd, Luogang District, China). In order to optimise for accuracy, known positive and negative samples were included as positive and negative controls in the assay.

Levels of alanine aminotransferase (ALT), Aspartate aminotransferase (AST), alkaline phosphatase (ALP), Total protein (TP), Albumin (ALB) and total bilirubin (TB) concentrations were determined by the use of an automated chemistry analyzer (Roche Hitachi Model 902).

### 2.4 Statistical Analysis

A power statistic was used to determine the number of samples required to at least detect a 20% difference in serum protein liver function markers with 80% power at a conventional level of alpha, 0.05. This design was used to reduce the likelihood of making type 1 and 2 errors or detecting an effect in the absence of any, and vice *versa*.

We used Minitab software version 18 for the interval plot in figure 1 and both Principal Component Analysis (PCA) and loading plot correlation analysis (Figure 4 and Table 4). An SPSS was used for forward regression analysis alongside with the PCA in Minitab software to ascertain the most important predictor variables that discriminates among disease groups. A Prism 10 software was used for both ROC analysis and Box plots showing Median/10-90 percentile, (Figure 2 and 3). An unpaired t-test (Mann-Whitney test) was used to make comparisons between two groups and ANOVA with multiple comparison testing, Bonferroni’s correction was used to make comparison for more than two groups. All statistical significance is determined at a conventional level of alpha (p<0.05). For some plots, designated signs were used on the plot/graph to explain different degree of statistical significance and such signs were explained in figures and table legends where applicable.

**Figure 1:**
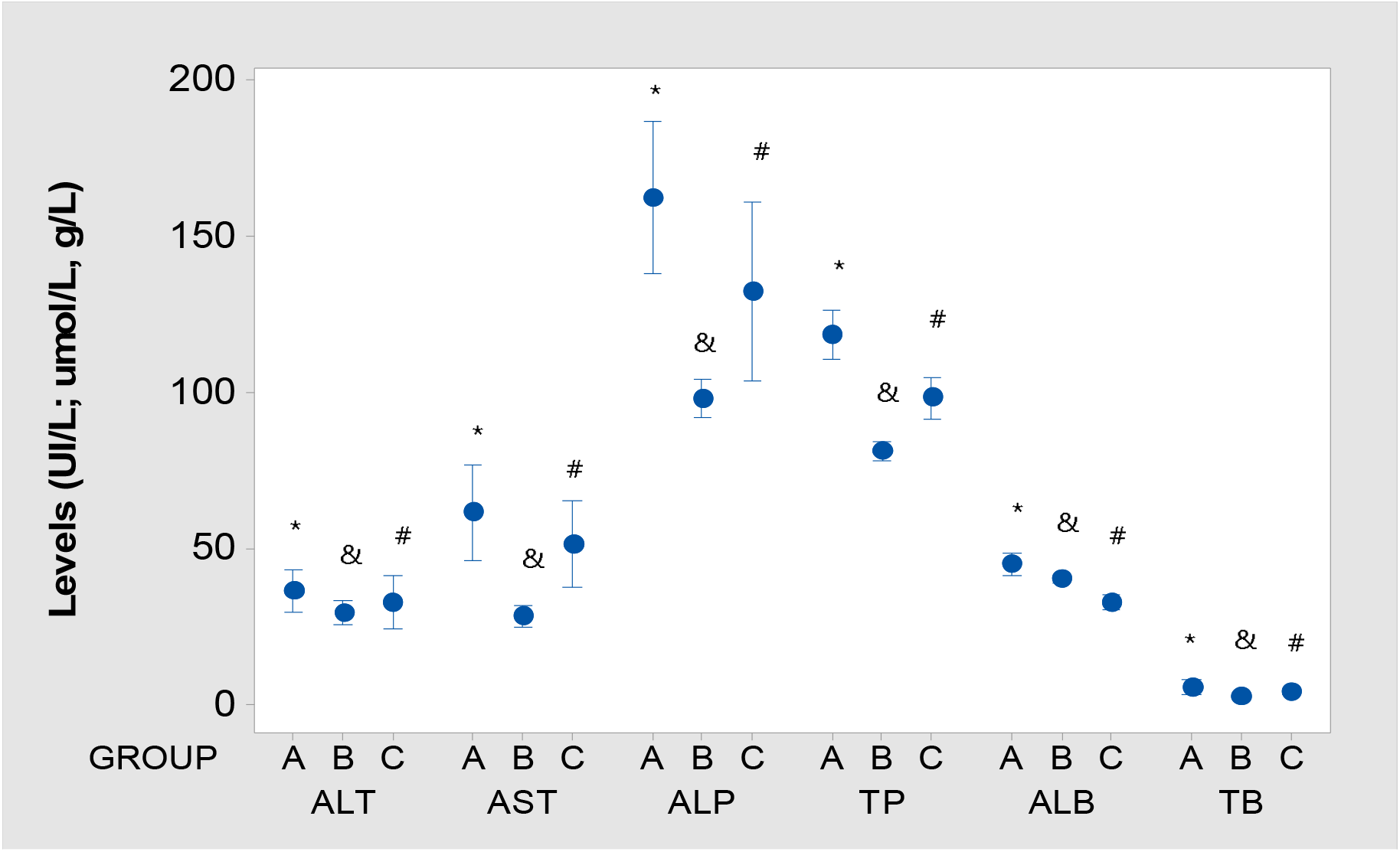
Serum protein liver function markers in study groups. **Notes**: A = HIV-1 seropositive patients receiving anti-retroviral drug; B = Healthy control group without HIV-1 and other overt aetiology; C = HIV-1 seropositive patients yet to commence anti-retroviral drug. * = Group A is statistically different (P < 0.001) to both group B and C; & = Group B is statistically different (P < 0.001) to both group A and C; # = Group C is statistically different (P < 0.001) to both group A and B. Values are presented as mean, interquartile range (IQR). N (number) = 150. **Abbreviations**: ALT, alanine aminotransferase; AST, aspartate transaminase; ALP, alkaline phosphatase; TP, total protein; ALB, albumin; TB, total bilirubin

**Figure 2:**
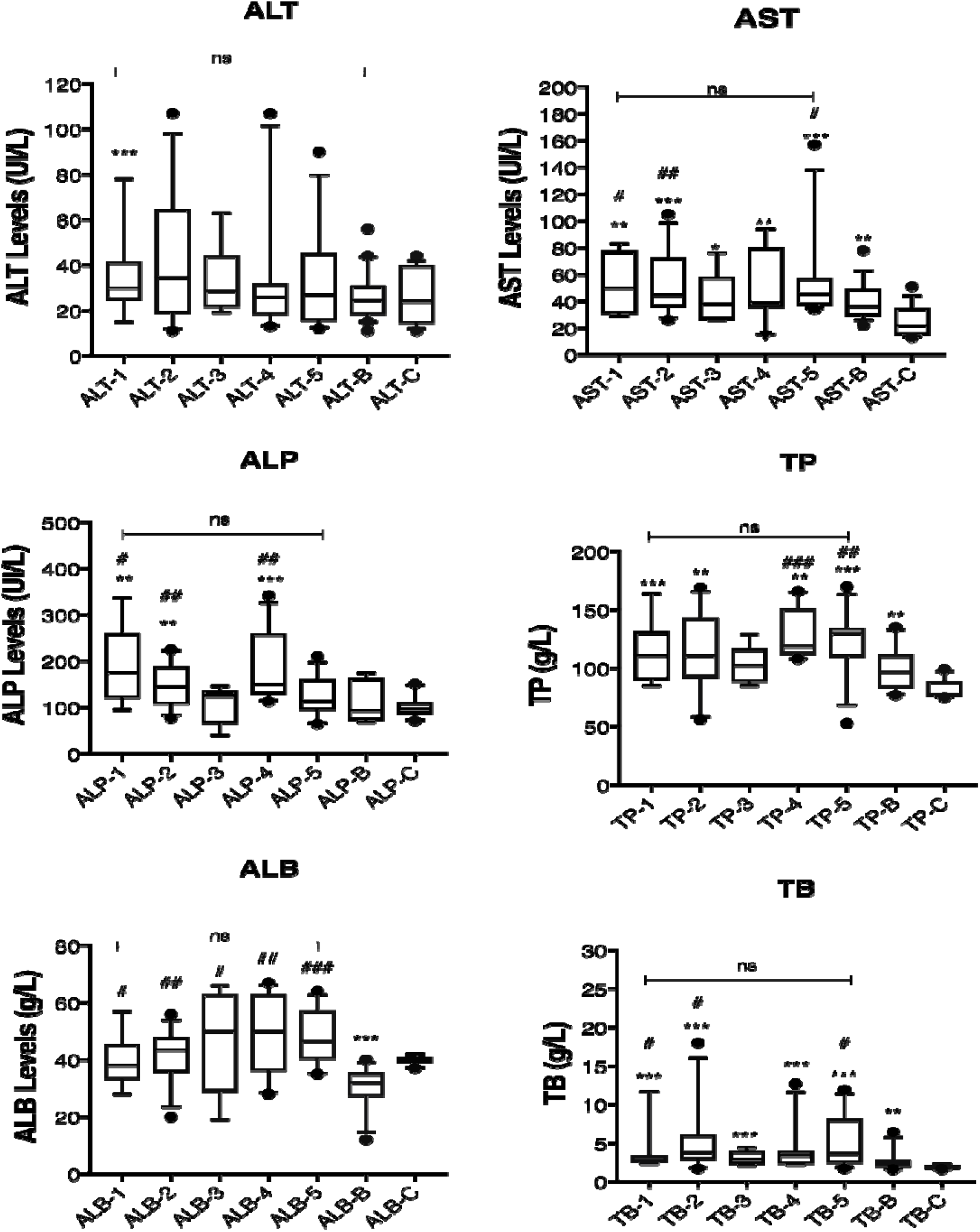
Liver function responses during a 5-year course of ART in comparison with levels in both the baseline and control groups. **Notes**: Values are presented as median, interquartile range (IQR). * = Statically different (* = 0.1; ** = 0.01 ; *** = 0.001) when compared to the healthy population, control group (ALT-C, AST-C, ALP-C, TP-C, ALB-C, TB-C). # = Statically different (# = 0.1; # # = 0.01; # # # = 0.001) when compared to the baseline data (ALT-B, AST-B, ALP-B, TP-B, ALB-B, TB-B). ANOVA with Bonferroni’s multiple comparison test (p<0.05) performed over groups in which line was drawn. Total number of data are 100, 50 on continuous ART for 5 years and with baseline data in addition to 50 in the control group. **Abbreviations**: ALT, alanine aminotransferase; AST, aspartate transaminase; ALP, alkaline phosphatase; TP, total protein; ALB, albumin; TB, total bilirubin; ns, not significant; The nomenclature, for example ALT-1, ALT-2 indicate average level within a given year. In this case, ALT-1 and ALT-2 represent ALT level in the first and second year of ART respectively. ALT-B, AST-B, ALP-B, TP-B, ALB-B and TB-B all represent baseline levels; ALT-C, AST-C, ALP-C, TP-C, ALB-C and TB-C all represent levels in the healthy control groups; g/L, gram per litre; UI/L, international units per litre.

**Figure 3.**
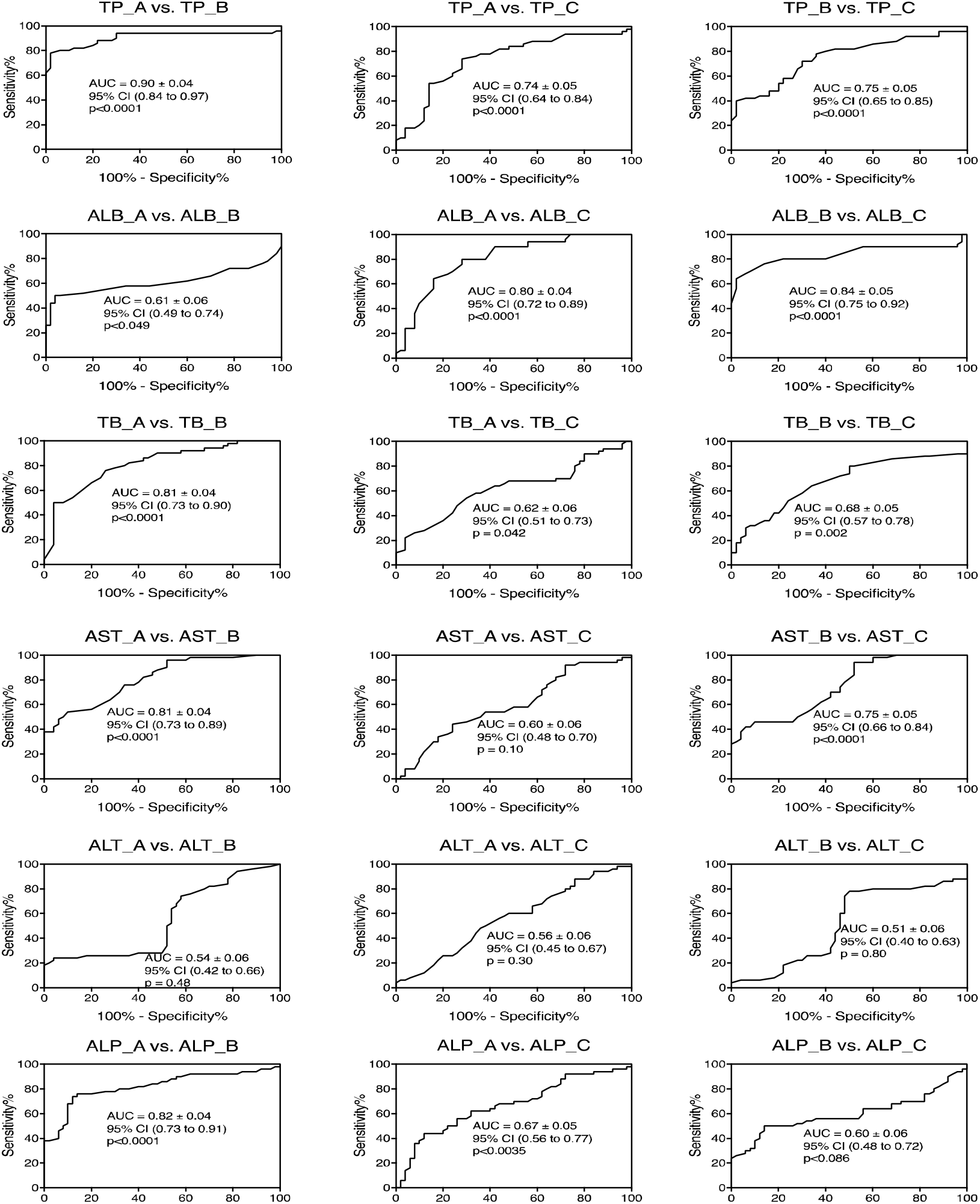
Serum protein liver function biomarkers discriminates between the study groups. Note: The Receiver Operating Characteristics (ROC) curve indicates that some of the markers well discriminate between the study groups. Graphs are shown as sensitivity against 100% - specificity. The AUC (Area Under the Curve), 95% Confidence Interval (CI) values and p-values are shown in the middle of each graph. AUC indicate how best marker discriminates between two groups. An AUC greater than 70% (0.7) with a significant p-value (p<0.05) is considered to be good at discriminating between groups. Abbreviations: ALT, alanine aminotransferase; AST, aspartate transaminase; ALP, alkaline phosphatase; TP, total protein; ALB, albumin; TB, total bilirubin. TP_A, total protein in Group A; TP-B, total protein in group B; vs, versus; A, Group A (Seropositive subjects receiving ART); B, Group B (HIV negative healthy population with no other overt aetiology and liver function problem and designated as the control group) and C, Group C (seropositive HIV subjects yet to commence ART).

**Figure 4.**
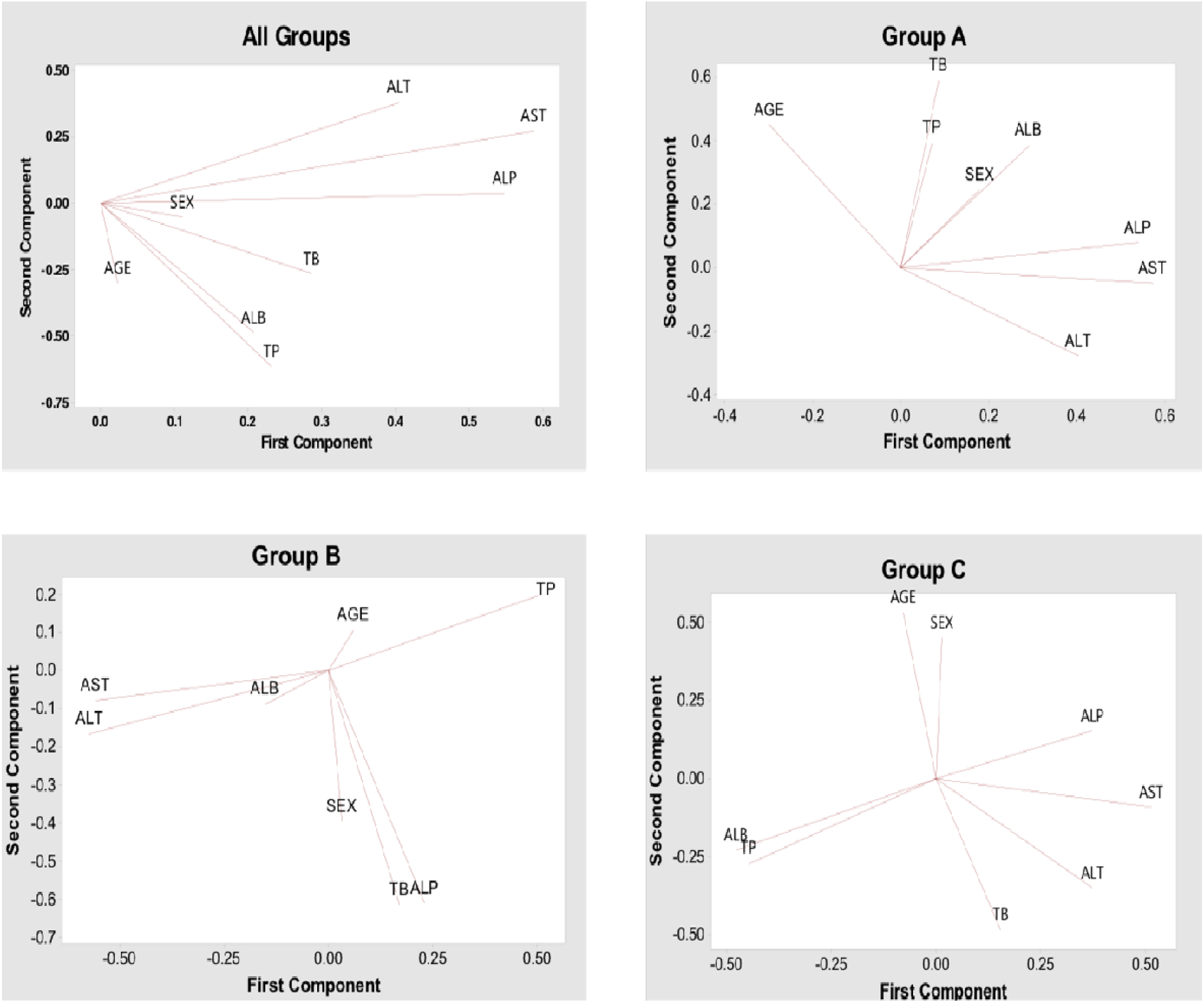
A loading plot showing the relationship among variables. **Note**: In the loading plot, the high positive correlation between two variables leads to two vectors that are very close to each other, forming a small angle. The non-correlation leads to two vectors out of phase by 90°, while the anti-correlation or negative correlation leads to two vectors that are out of phase by 180°. Seropositive HIV-1 subjects on ART (Group A); healthy population control group with no overt aetiology (group B) and seropositive HIV-1 subjects yet to commence ART (group C). Plots reveals only the first and second components. **Abbreviations**: ALT, alanine aminotransferase; AST, aspartate transaminase; ALP, alkaline phosphatase; TP total protein; ALB, albumin; TB, total bilirubin.

## 3 Result

### 3.1 Seropositive HIV-1 patients with or without ART exhibits different levels of serum markers to the control group

Seropositive HIV-1 patients on ART (Group A) have significantly higher (p<0.001) levels of serum markers, ALT, AST, ALP, TP, ALB and TB than both seropositive HIV-1 yet to commence anti-retroviral therapy (Group C) and the healthy non-HIV positive, the group B (Figure 1). The group B exhibits significantly lower levels (p<0.001) of serum makers compared to group A and C except for the ALB levels which is higher than the group C (Figure 1). The levels of these proteins are independent of age and sex (Data not shown) in all the clinical groups.

### 3.2 The duration of Anti-Retroviral Therapy (ART) is not associated with significant changes in serum markers

The levels of serum markers were not statistically different all through the 1st to 5th year duration of ART, (Figure 2). However, while serum marker at some point during the course of ART showed to be significantly higher when compared with baseline level, levels at other time points do not show to be different to baseline level (Figure 2). Unlike other markers, the ALT level remained stable all through the course of ART and do not show any significant difference to baseline level (Figure 2).

The baseline level of ALB is significantly lower compared to the levels in the control (healthy population) and increases at initiation of ART and slightly during the course of the therapy (Figure 2). The baseline levels in both the ALT and ALP were not different to levels in the healthy control group unlike the ALB, AST, TP and TB levels (Figure 2).

### 3.3 Serum protein discriminate among seropositive patients with and without anti-retroviral therapy and healthy HIV negative subjects

Having observed that there are distinct differences in mean in the interval plots (Figure 1), we analysed the data for how well these markers could discriminate or separate the groups using a Receiver Operating Characteristic (ROC) curve’s sensitivity versus 100%- specificity (Figure 3). The level of TP showed to be very good at discriminating among all the groups: seropositive HIV-1 subjects on ART (Group A), seropositive HIV-1 subjects yet to commence ART (Group C) and non-HIV positive healthy group, Group B (Figure 3). The levels of ALB and TP are very good in discriminating group A from group C (Figure 3). Furthermore, the ALB levels further show to best separate group B from group C (Figure 3).

Overall, a forward regression analysis indicated that both ALB and TP levels are the most important predictor variables in the study groups and control up to 27.2% of the observed differences among the groups (Table 2), ALB being the strongest, r^2^ = 0.249; p=0.000 (Table 2) predictor. AST, ALP, TP and ALB variables contributed to the first and second Principal Components (PC) in PCA analysis than other variables (Table 3).

**Table 2.**
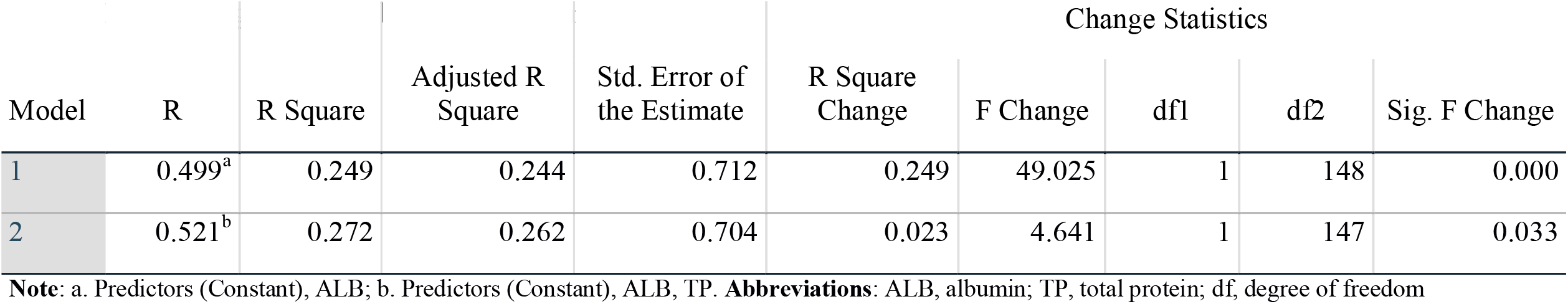
Forward regression analysis model summary.

**Table 3.** Eigenvectors indicating important variable in the principal components.

| Variable | PC1 | PC2 | PC3 | PC4 | PC5 | PC6 |
| --- | --- | --- | --- | --- | --- | --- |
| ALT | 0.413 | -0.357 | 0.432 | 0.462 | 0.408 | 0.369 |
| AST | 0.591 | -0.281 | -0.152 | 0.021 | -0.040 | -0.739 |
| ALP | 0.547 | -0.053 | -0.435 | -0.244 | -0.368 | 0.560 |
| TP | 0.238 | 0.616 | -0.351 | 0.010 | 0.664 | -0.007 |
| ALB | 0.206 | 0.585 | 0.227 | 0.552 | -0.506 | -0.061 |
| TB | 0.286 | 0.263 | 0.653 | -0.650 | 0.008 | -0.025 |

**Notes:** AST, ALP, TP and ALB variables contributed to the first and second Principal Components (PC) in PCA analysis than other variables
**Abbreviations:** ALT, alanine aminotransferase; AST, aspartate transaminase; ALP, alkaline phosphatase; TP, total protein; ALB, albumin; TB, total bilirubin; PC, principal components

**Table 4:**
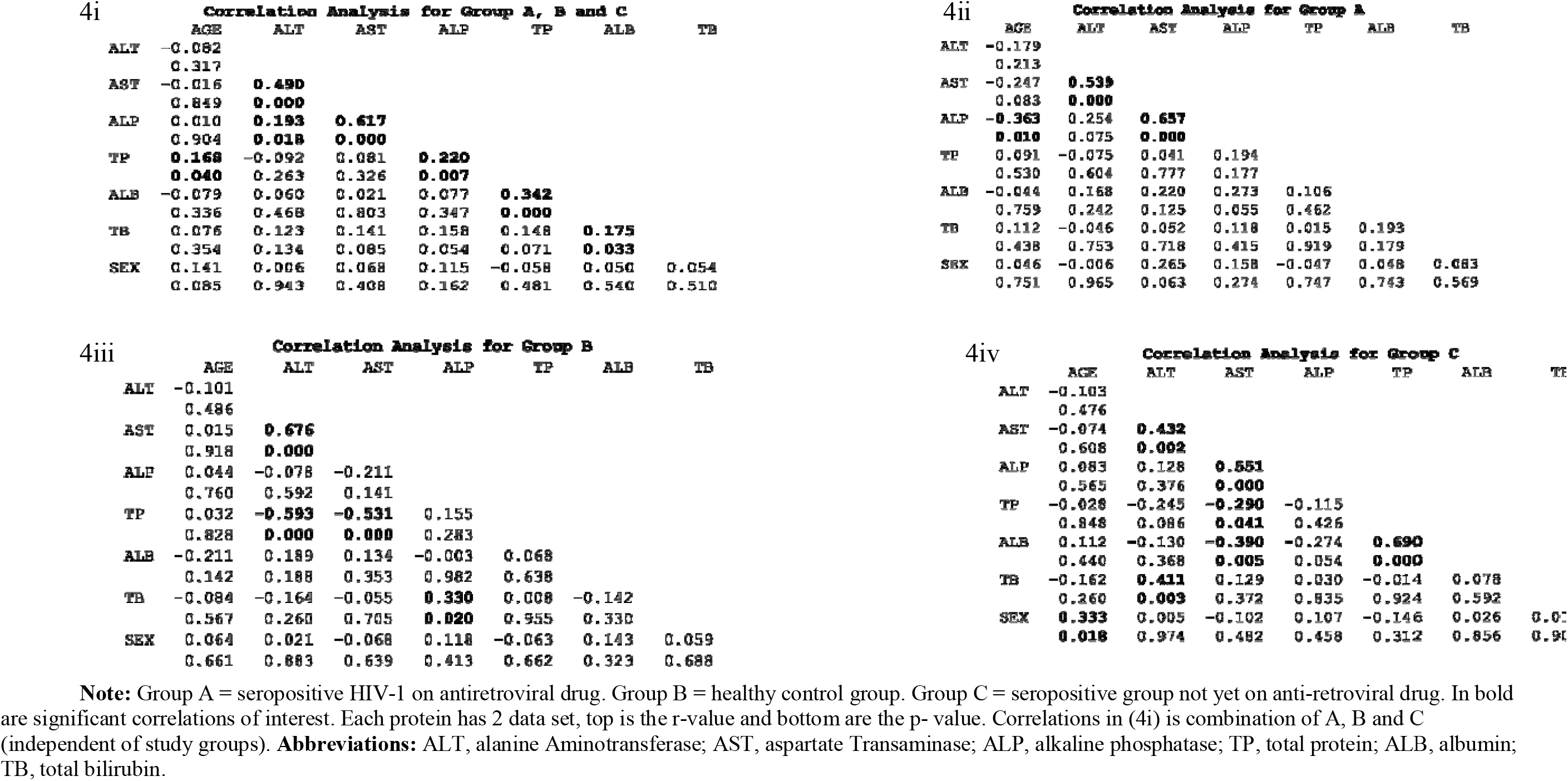
Correlation tables for liver function protein in the different study groups.

### 3.4 Serum protein correlates differently in the healthy subjects and seropositive patients with and without ART

The correlative distribution pattern (loading plot) of the serum proteins differs in the different study groups (Figure 4i-iv, Tables 4i-iv). The observed pattern in group A (Figure 4ii; Table 4ii) and group C (Figure 4iv; Table 4iv) could be assumed to be associated with their respective drug effect and/or pathophysiology while the correlation pattern in group B (Figure 3iii; Table 3iii) could be assumed or considered to be the normal. A significant positive correlation between TB and ALT, ALB and TP and a significant negative correlation between ALB and AST differentiated group C (Figure 4iv; Table 4iv) to group A (Figure 4ii, Table 4ii). A significant negative/inverse correlation between TP and ALT and a positive correlation between TB and ALP are unique to healthy subjects, group B (Figure 4iii and Table 4iii). TP and age, ALP and ALT, ALP and TP, TB and ALB all correlated independent of the study groups (Table 4i).

## 4.0 Discussion

Serum proteins like ALT, AST, ALP, ALB, TP and TB are widely used as indicators in evaluation of patients for hepatic dysfunction and are widely referred to as liver function protein markers and are often shown to reflect the pathological conditions of other internal organs like heart and kidney as well (Gowda et al., 2009). The assessment of hepatic effect of ART among HIV patients could be a daunting challenge owing to multiple cofounding factors. A major strength in this study is the well clinically defined 5 years retrospective sample data set from HIV-1 mono-infected subject, including both baseline and control data and without cofounding factors.

The significantly higher level of some serum protein liver function markers in seropositive group yet to commence ART as compared to the control group possibly suggests a higher degree of liver injury owing to the infection itself. Previous report showed that more damage is caused by the infection than antiretroviral drug which is thought to be less active in the liver (Crane, Iser, & Lewin, 2012; Ganesan, Poluektova, Kharbanda, & Osna, 2018; Jones & Núñez, 2012; Lucien et al., 2010; Osakunor et al., 2015).

The findings indicate that the increasing duration of ART does not cause a significant change in levels of serum protein liver function markers nor indicative of increasing liver damage as the levels of these liver function markers do not show any significant change all through the 5-year period of ART.

Both the ALP and AST levels show a slight decrease in median from the 1^st^ to 3rd year of ART therapy. This is in line with a previously reported decrease in liver function markers after the first year with continuous ART use in HIV subjects without co-infection with HBV and HCV (Qin et al., 2019).

Low ALB level in seropositive subjects yet to commence ART and its high level at ART initiation likely indicate both the role of ALB in the pathological process and a possible mechanism of ART. Furthermore, high level of ALB might be important in the recovery process and ART might work towards increasing ALB level as a possible mechanism of action. Level of ALB has been a promising prognostic marker in HIV patients with or without ART (Dirajlal-Fargo et al., 2018; Feldman et al., 2000; Olawumi & Olatunji, 2006; Sundaram et al., 2009).

The levels of some serum proteins show to be discriminatory among seropositive subjects receiving ART, those without ART and the healthy control groups: while the level of total protein (TP) showed to be excellent in discriminating between seropositive HIV-1 subjects on ART and healthy subjects, levels of albumin (ALB) is discriminatory among seropositive HIV-1 subjects on ART; seropositive subjects yet to commence ART and the healthy control group (Figure 3). The observation here is further supported by previous report that HIV mono-infected patient has both elevated levels of TP and ALB unlike in HIV co-infection with hepatitis where only elevated levels of ALB was reported (Serpa, Haque, Valayam, Breaux, & Rodriguez-Barradas, 2010). These proteins are therefore promising prognostic biomarkers in mono-infected HIV-1 subjects with or without ART: more so that the observed differences in levels are independent of both sex and age as observed in this cohort and as previously reported (Lucien et al., 2010). Both TP and ALB levels might be tightly linked to the pathophysiology of the disease in the seropositive HIV-1 subjects receiving or yet to commence ART and their roles need to be further studied. Furthermore, the loading plots pattern and correlation analysis among these proteins further suggest differences in pathophysiology between the seropositive subjects with or without ART and these proteins interactions in pathological condition also requires further studies.

The observed normal level of liver aminotransferase enzyme (ALT) from the 2^nd^ year of ART initiation could be indicative of low inflammatory response as previously reported (Osakunor et al., 2015). While liver damage is thought to persist is some subjects with normal ALT level (Uberti-Foppa C1, De Bona A, Galli L, Sitia G, Gallotta G, Sagnelli C, Paties C, 2006), others have reported a positive prognosis when transaminase level is restored to normal (Segamwenge & Bernard, 2018). Therefore, the normal ALT level observed from the second years of ART initiation may indicate positive prognosis (Segamwenge & Bernard, 2018). However, the conflicting report on the role of ALT in HIV patients receiving ART need to be further investigated.

This well-defined study is important in the management of HIV patients in resource and skill limited communities: both the ALB and TP levels are promising biomarkers in differentiating among HIV-1 positive subjects receiving or yet to commence ART and could be important in HIV management in poor resource district hospitals. The study also shed some light on the pathophysiology of HIV-1 infection in subjects with or without ART

## 5.0 Conclusion and recommendation

There is no significant increase in the levels of serum protein biomarkers of liver function with continuous use of ART beyond the levels observed on initiation of ART. Hence, hepatic dysfunction with continuous use of ART is unlikely. Both the ALB and TP serum level may be important in the management of HIV patients on anti-retroviral therapy in resource limited communities.

This report looks at ART in general rather than assessing the effect of each anti-retroviral drug/regimen. It will be important to look into the effect of each antiretroviral drug/regimen on levels of serum protein markers of liver function in future studies.

## Data Availability

Data available on demand

## Acknowledgement

The authors thank the patients whose data were used in this study.

## Declaration

Author(s) declare that there are no source of funding and no conflicts of interest in this work.

## Authors’ contribution

AES, MH and SEA designed the study. SEA, and MH supervised the project. AES, MH, TS and YBN carried out the experiment and data collection. SEA, EEG, SS, DB and JIO analysed the data. SEA, DB, TS, EEG, EFT and DAM revise the manuscript and performed the interpretation of the statistical analysis. AES, SEA, DAM, DB, JIO and SS wrote and proof read the manuscript. All authors have read and agreed on the final version of this manuscript.

